# Antidepressant and antipsychotic drug prescribing and complications of diabetes: a systematic review of observational studies

**DOI:** 10.1101/2023.01.27.23285097

**Authors:** Charlotte R L Greene, Hanna Ward-Penny, Marianna F Ioannou, Sarah H Wild, Honghan Wu, Daniel J Smith, Caroline A Jackson

**Author notes:** Corresponding author: Dr Caroline Jackson.

## Abstract

**Aims:** Psychotropic medication may be associated with adverse effects, particularly in people with diabetes. We conducted a systematic review of observational studies investigating the association between antidepressant or antipsychotic drug prescribing and diabetes outcomes.

**Methods:** We systematically searched PubMed, EMBASE, and PsycINFO to 15^th^ August 2022 to identify eligible studies. We used the Newcastle-Ottawa scale to assess study quality and performed a narrative synthesis.

**Results:** We included 18 studies, 14 reporting on antidepressants and four on antipsychotics. There were 11 cohort studies, one self-controlled before and after study, two case-control studies, and four cross-sectional studies, of variable quality and highly heterogeneous in terms of study population, exposure definition and outcome analysed. Antidepressant prescribing may be associated with increased risk of macrovascular outcomes, whilst evidence on antidepressant and antipsychotic prescribing and glycaemic control was mixed. Few studies reported on microvascular complications and cardiometabolic factors other than glycaemic control and just one study reported on antipsychotics and diabetes complications.

**Conclusions:** There has been little study of antidepressant and antipsychotic drug prescribing in relation to diabetes outcomes. Further, more methodologically robust, research is needed to inform and enhance antidepressant and antipsychotic drug prescribing and monitoring practices in people with diabetes.

## 1.1 Introduction

Antidepressant and antipsychotic drugs are common psychotropic medications used in treating mental illness such as major depression, bipolar disorder, and schizophrenia [1, 2], with prescribing having increased in high income countries in recent years [3, 4]. This may be partly driven by longer treatment duration, but also by more frequent use, including for other indications (such as agitation and aggression in autism spectrum disorder and dementia [5] and chronic pain, migraines, and insomnia [6]). This increased use is of psychotropic drugs is of particular concern given that they appear to be associated with risk factors for cardiovascular disease and type 2 diabetes such as obesity, insulin resistance, and dyslipidaemia [7, 8], as well as major cardiovascular events such as coronary heart disease and stroke [9, 10].

These adverse effects must be considered in the context of their use among people with diabetes, including the bidirectional links between mental illness and diabetes [11] and the use of tricyclic antidepressants (TCA) to treat neuropathic pain occurring as a complication of diabetes [12]. Previous reviews have summarised evidence from experimental studies on the effect of antidepressants on cardiometabolic risk factors in people with diabetes [13, 14]. Although these reviews concluded that some antidepressant subtypes may be associated with improved glycaemic control, the overall evidence was inconclusive, in part due to low study quality, small sample sizes, and insufficient follow-up to adequately investigate longer-term outcomes such as vascular morbidity or mortality.

To our knowledge there is no previous systematic review of observational studies reporting the association between antidepressant or antipsychotic drugs and diabetes outcomes. We therefore conducted a systematic review of observational studies in people with diabetes that examined the association between antidepressant or antipsychotic drug prescribing and clinical complications of diabetes (including cardiovascular and microvascular morbidity), mortality, or cardiometabolic risk factors.

## 2.1 Materials and methods

This review is reported in accordance with the PRISMA guidelines. The review protocol was not registered

### 1.2.1 Data sources and searches

We searched PubMed, EMBASE, and PsycINFO from point of inception to August 15^th^ 2022 using a comprehensive electronic search strategy (Supplemental appendix 1).

### 1.2.2 Study selection

We included cohort, case-control, self-controlled or cross-sectional studies conducted among adults with type 2 diabetes that examined the association between prescribing of any antidepressant and/or antipsychotic prescribing versus no prescribing in relation to diabetes outcomes (macrovascular or microvascular complications; all-cause mortality; or cardiometabolic risk factors [glycaemic control; blood pressure; or lipid levels]). See Supplemental Table 1 for a detailed summary of the inclusion criteria. We did not limit our search to studies published in the English language, but ultimately only included English language articles.

Two reviewers (CRLG and HWP or MFI and CAJ) independently screened titles and abstracts and the full-text of potentially eligible articles and extracted information on study and population characteristics, exposure and outcome/case definitions, statistical methods, and results from included articles. Disagreements about suitability for inclusion or data extracted were resolved through discussion with a third reviewer (CAJ or SHW).

### 1.2.3 Quality assessment

Two reviewers (CRLG and HWP or MFI and CAJ) assessed methodological study quality using the Newcastle-Ottawa scale [15]. This assesses the quality of observational studies across eight items within three domains: (i) the selection of the study groups; (ii) the comparability of the groups; (iii) the ascertainment of either the outcome or exposure of interest, where relevant.

### 1.2.4 Data synthesis and analysis

We performed a narrative synthesis since substantial clinical and methodological heterogeneity between studies precluded meta-analysis.

## 3.0 RESULTS

### 3.1 Study characteristics

Our search yielded 11,216 articles, with 19 eligible articles representing 18 studies ultimately included (Figure 1).

**Figure 1.**
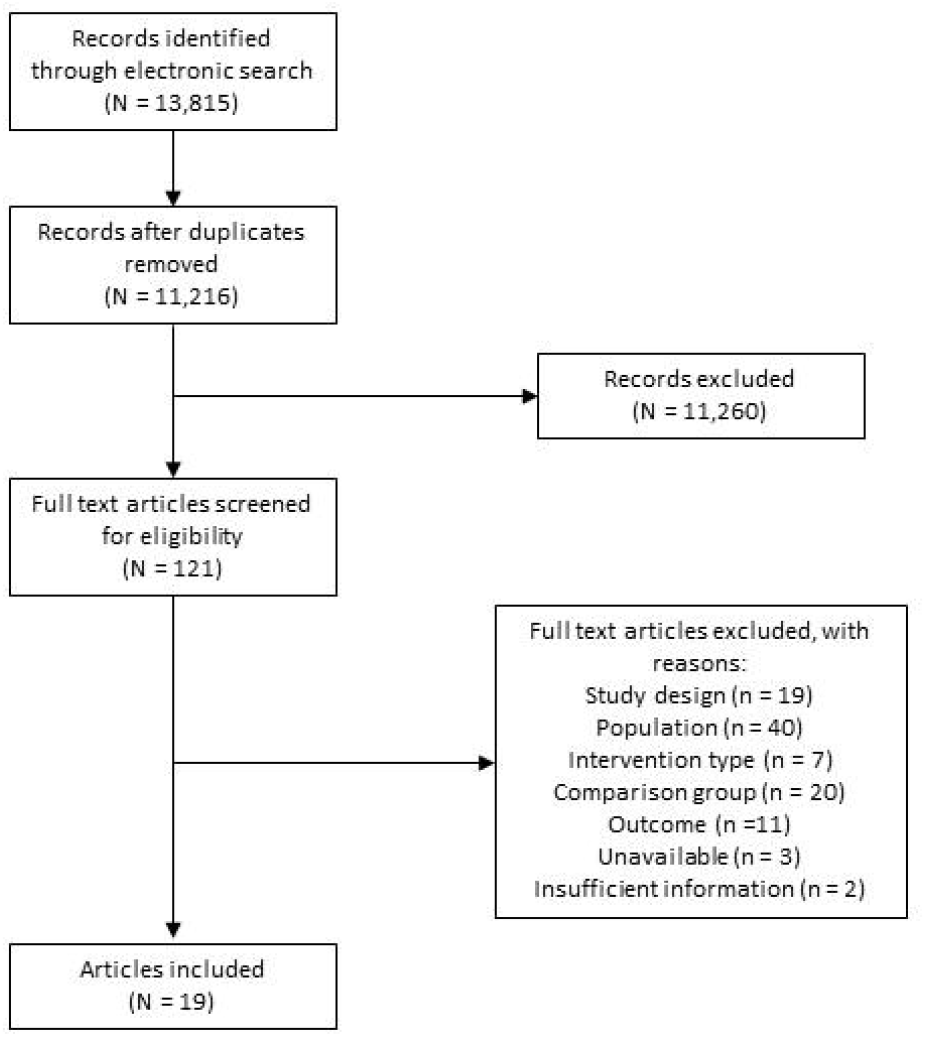
Study selection flow diagram

Study characteristics are described in Table 1. Details of the antidepressant and antipsychotic drugs included with each study are given in Supplemental Table 2. Eleven studies were cohort [16-26], one was a self-controlled before and after study [27], two were case-control [28, 29], and four were cross-sectional studies [30-33]. Two studies included overlapping study populations [21, 27], but adopted different study designs to address a similar question and so both were included in the narrative synthesis. There was considerable heterogeneity between studies in terms of study population, exposure definitions and outcomes analysed. All studies were from high-income settings. Study populations were most frequently derived from people with newly diagnosed diabetes in the general population. Two studies included a study population with comorbid mental illness and diabetes (one with depression [25] and one with schizophrenia [24]).

**Table 1:**
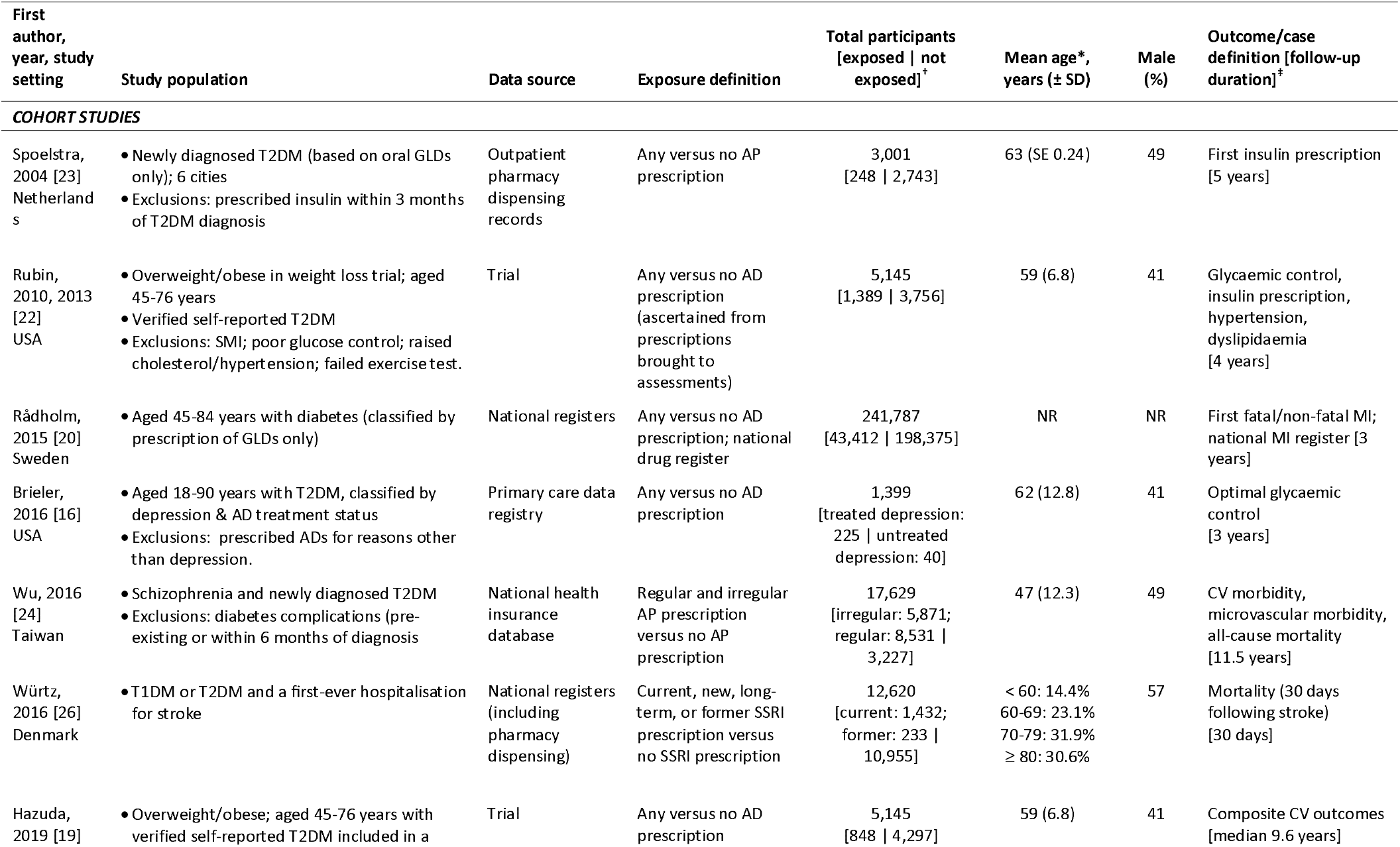
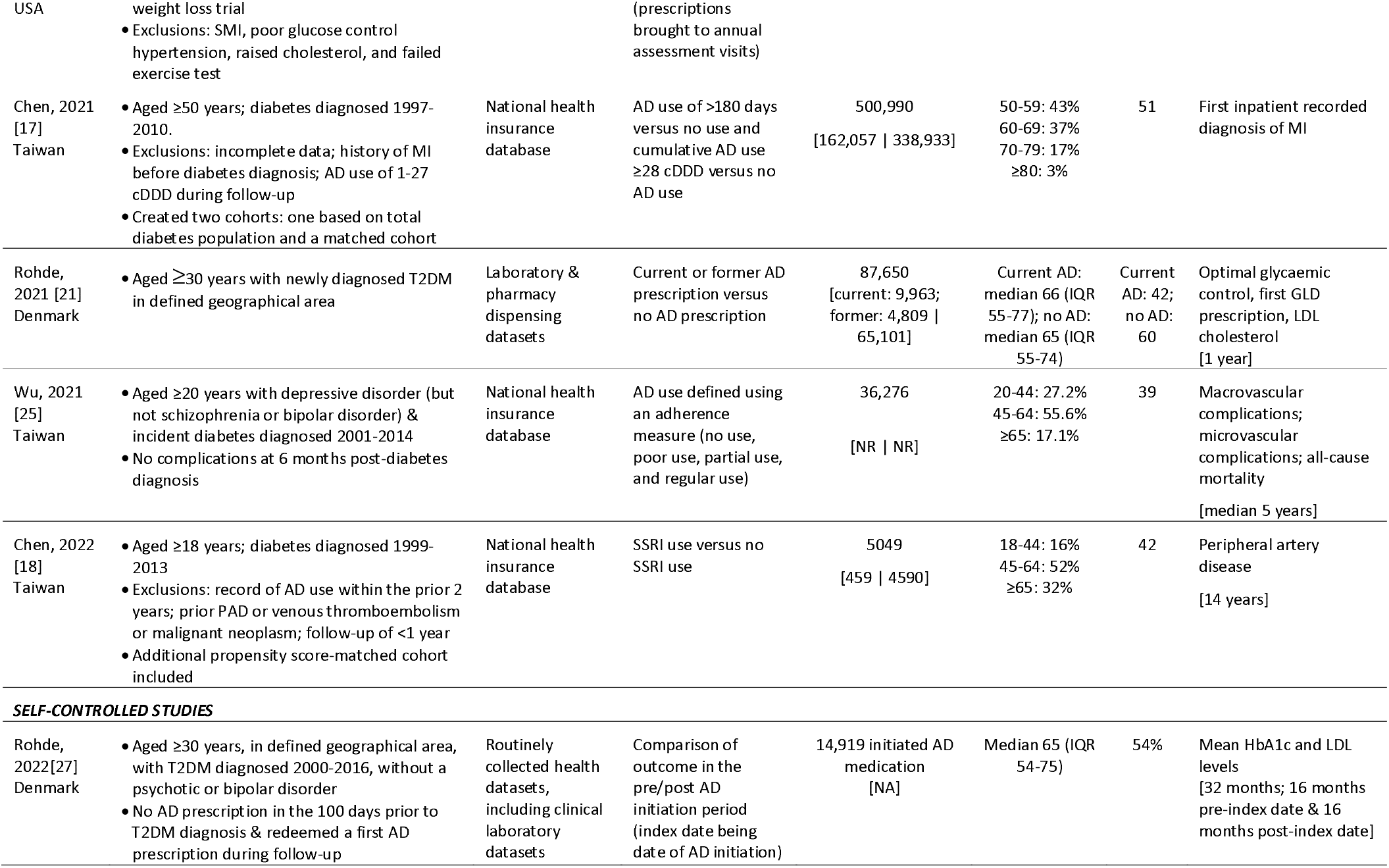
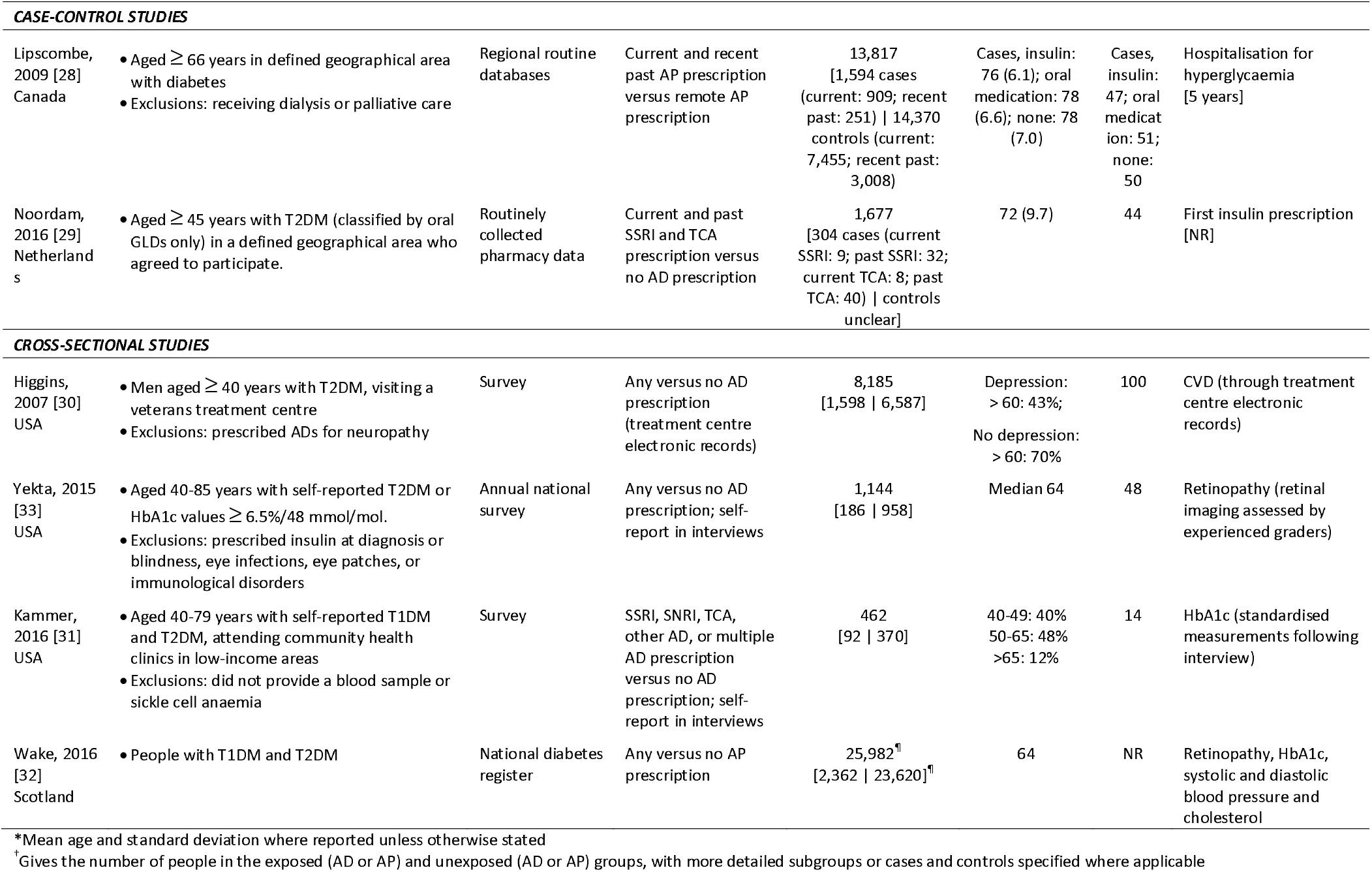
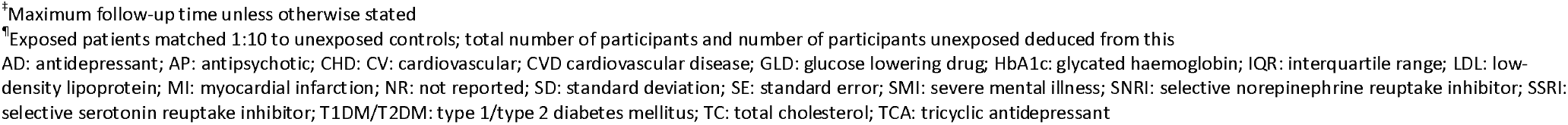
Characteristics of included studies

### 3.2 Quality assessment

Methodological study quality ranged from three to eight out of nine stars (Supplemental Table 3). Concerns arising in some studies included: poor comparability due to insufficient adjustment for relevant confounders including confounding by indication; use of self-report for exposure and/or outcome assessment; poor representativeness of the population; low precision due to small sample sizes; and no description of the response rate and/or attrition (where applicable).

### 3.3 Antidepressant prescribing and diabetes outcomes

#### 3.3.1 Diabetes complications

Of 14 studies reporting on antidepressant prescribing, 8 reported associations with complications (largely macrovascular) of diabetes, comprising 6 cohort studies [17-20, 25, 26] and two cross-sectional studies [30, 33] (Table 2 and Supplementary Table 4a). Outcomes were heterogeneous and findings were mixed.

Receipt of antidepressant prescriptions, in comparison to no record of anti-depressant prescribing, was associated with higher crude incidence of myocardial infarction (MI) [20] and of a composite macrovascular outcome [25]. However, lack of adjustment for age, sex and other factors that may differ between groups limits conclusions from these studies. A third cohort study found that selective serotonin reuptake inhibitor (SSRI) antidepressant prescribing was associated with increased risk of 30-day post-stroke mortality [26], with the excess risk greatest for people whose SSRI prescription was initiated shortly before stroke. A cross-sectional study of older men of low-income from one treatment centre reported greater odds of a composite cardiovascular morbidity outcome and of MI specifically in crude analyses without control of confounding factors [30]. In contrast, a cohort study with a median of 9.6 years follow-up derived from post-hoc analyses of a clinical trial for a weight loss intervention [19] reported that antidepressant prescribing was associated with reduced cardiovascular morbidity or mortality in women, but not men. Risk of peripheral artery disease was reported to be similar in those with diabetes prescribed versus not prescribed antidepressants [18]. Finally, the only cohort study to describe microvascular complications reported higher crude incidence of a composite microvascular outcome in people prescribed anti-depressants among a Taiwanese cohort of people newly treated for diabetes [25], whilst a small cross-sectional study of people with a diagnosis of diabetes or with HbA1c 6.5% who took part in the National Health and Nutrition Examination Survey in the US found that self-reported antidepressant was associated with lower odds of retinopathy compared to people not reporting anti-depressant use after adjusting for sociodemographic and clinical characteristics [33] (Table 2). Supplemental Table 5a includes details of additional results reported in some studies, including investigations of variations of composite outcomes, interactions and associations by antidepressant drug subtypes, with findings generally consistent with primary results.

**Table 2:**
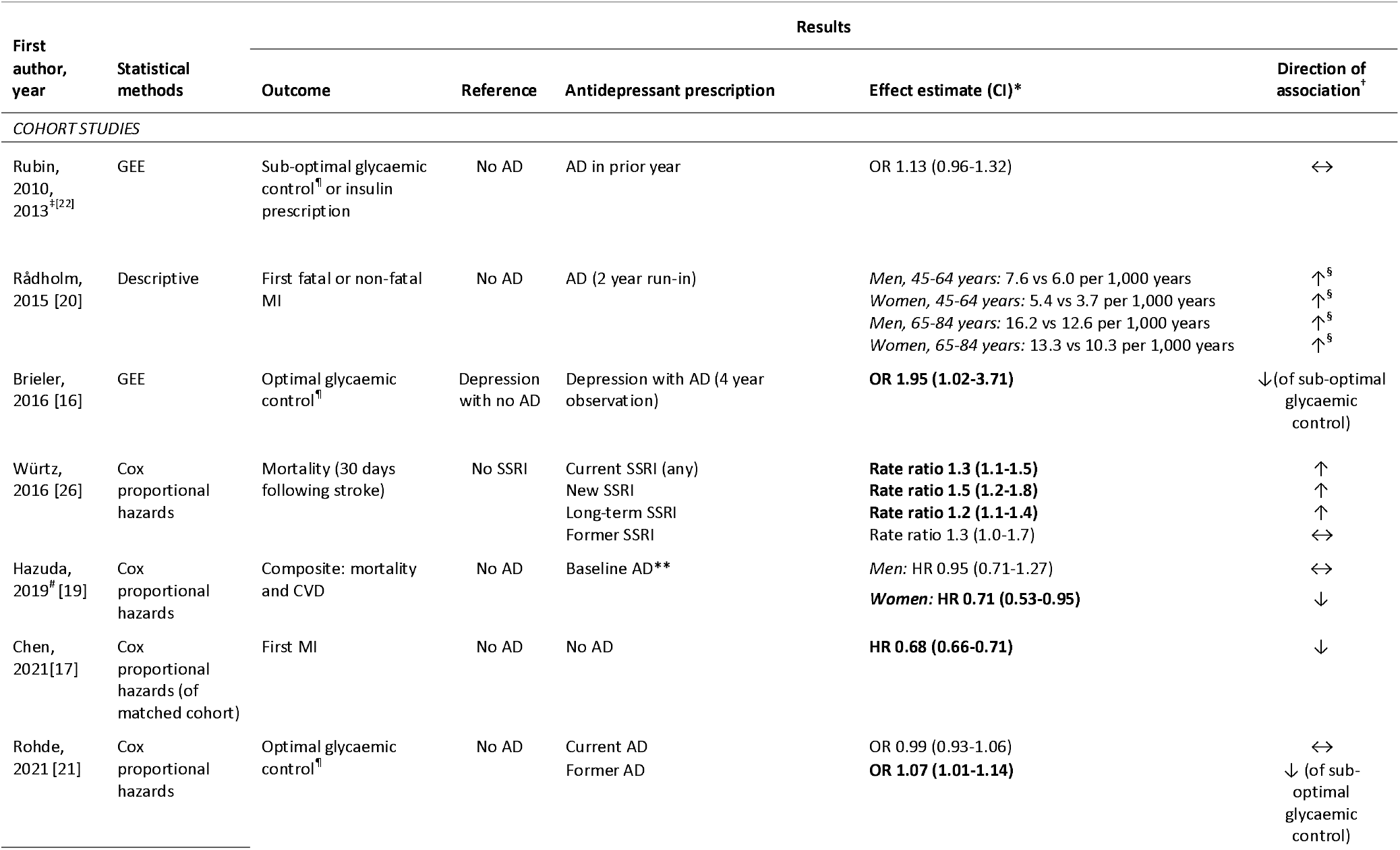
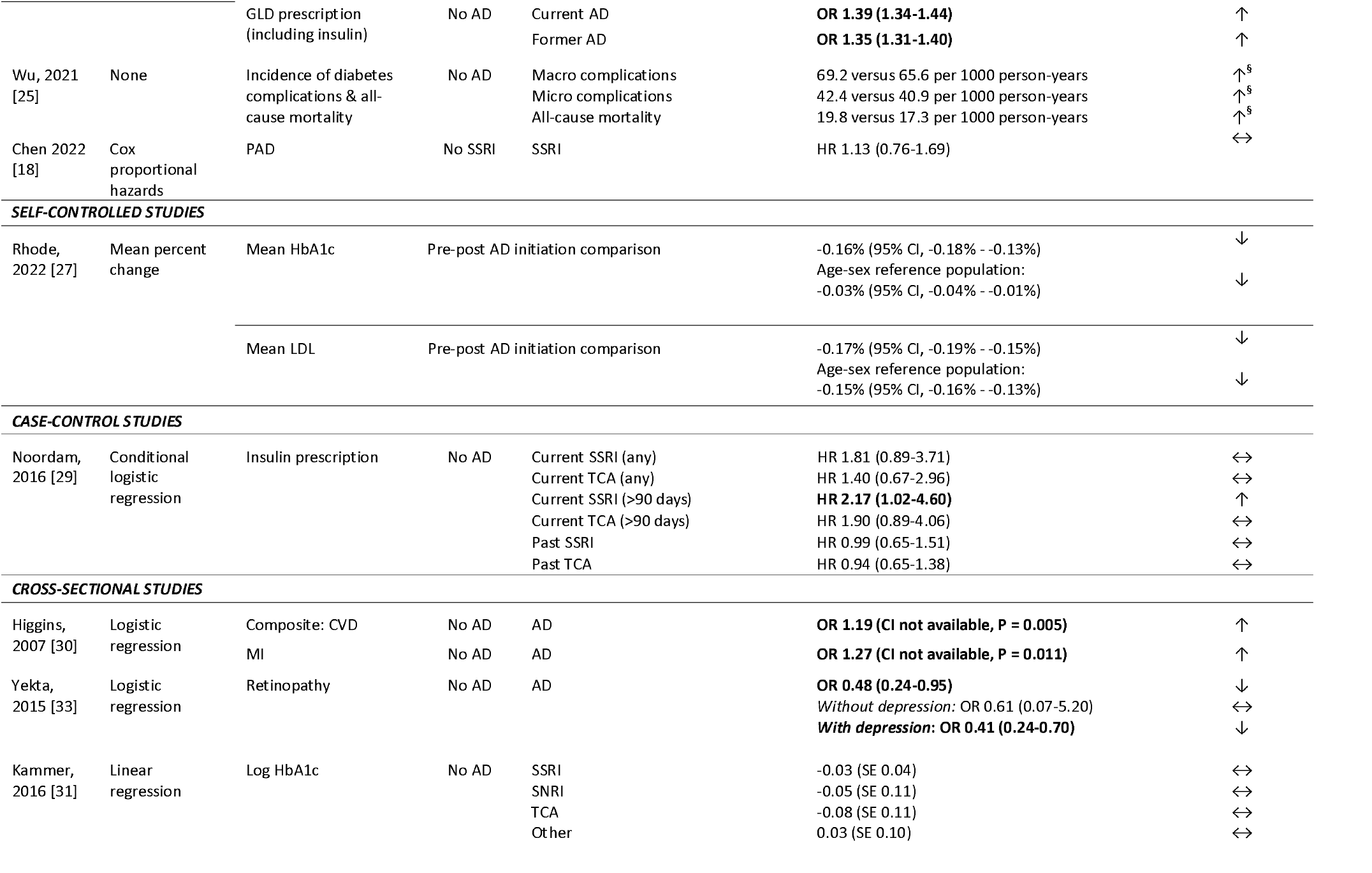
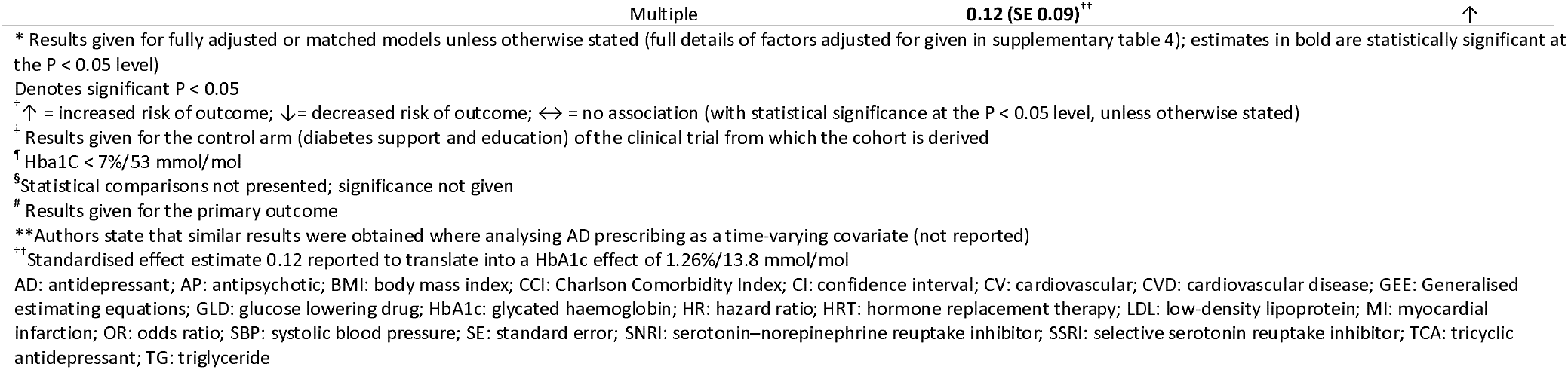
Results of observational studies reporting on the association between antidepressant drug prescribing and outcomes in people with diabetes

#### 3.3.2 Glycaemic control

Six studies reported on antidepressant prescribing in relation to glycaemic control [16, 21, 22, 27, 29, 31], with mixed findings. Of the three cohort studies, two reported no statistically significant difference in the association between antidepressant prescribing and optimal glycaemic control [21, 22]. The third study reported that antidepressant use was associated with lower risk of sub-optimal glycaemic control [16]. Interestingly, a pre-post study reported lower HbA1c levels following antidepressant medication initiation [27]. One cohort study also reported that receipt of antidepressants was associated with increased odds of prescribing of glucose lowering drugs (GLDs) [21]. Similarly, a case-control study found that among people with newly diagnosed diabetes, current receipt of SSRI antidepressant prescriptions was associated with a more than two-fold increased risk of insulin prescription [29]. Similar associations were observed for other antidepressant classes and for other durations, with no associations observed for past prescriptions (Table 2). A cross-sectional study among people from low-income areas found that antidepressant prescribing from multiple classes was associated with higher values of HbA1c, after adjustment for depression severity [31], whereas there was no association between individual antidepressant class prescribing and HbA1c levels.

#### 3.3.3 Lipid profile and lipid-lowering treatment

One cohort study found that antidepressant prescribing was associated with abnormal lipid profiles or receipt of lipid-lowering medication [22], whereas a second reported that antidepressant prescribing may have a small protective effect on cholesterol levels [21], with findings broadly consistent by prescription timing and across antidepressant subtypes (Supplemental Table 5a).

### 3.4 Antipsychotic prescribing and diabetes outcomes 3.4.1 Diabetes complications

Just four studies reported on antipsychotic prescribing in relation to diabetes outcomes. The two studies that investigated the association between antipsychotic prescribing and clinical complications of diabetes used population-based national registers [24, 32]. A cohort study of people with schizophrenia found that, compared to no antipsychotic prescribing, regular antipsychotic prescribing was associated with a 20% and 27% reduced risk of cardiovascular morbidity, and all-cause mortality, respectively [24] (Table 3). There was no clear evidence of an association between antipsychotics and microvascular complications. When stratifying by metabolic risk of antipsychotics, the authors found that drugs considered to have an intermediate or high metabolic risk were associated with a significantly reduced risk of all complications including cardiovascular and microvascular morbidity (Supplemental Table 5b).

**Table 3:**
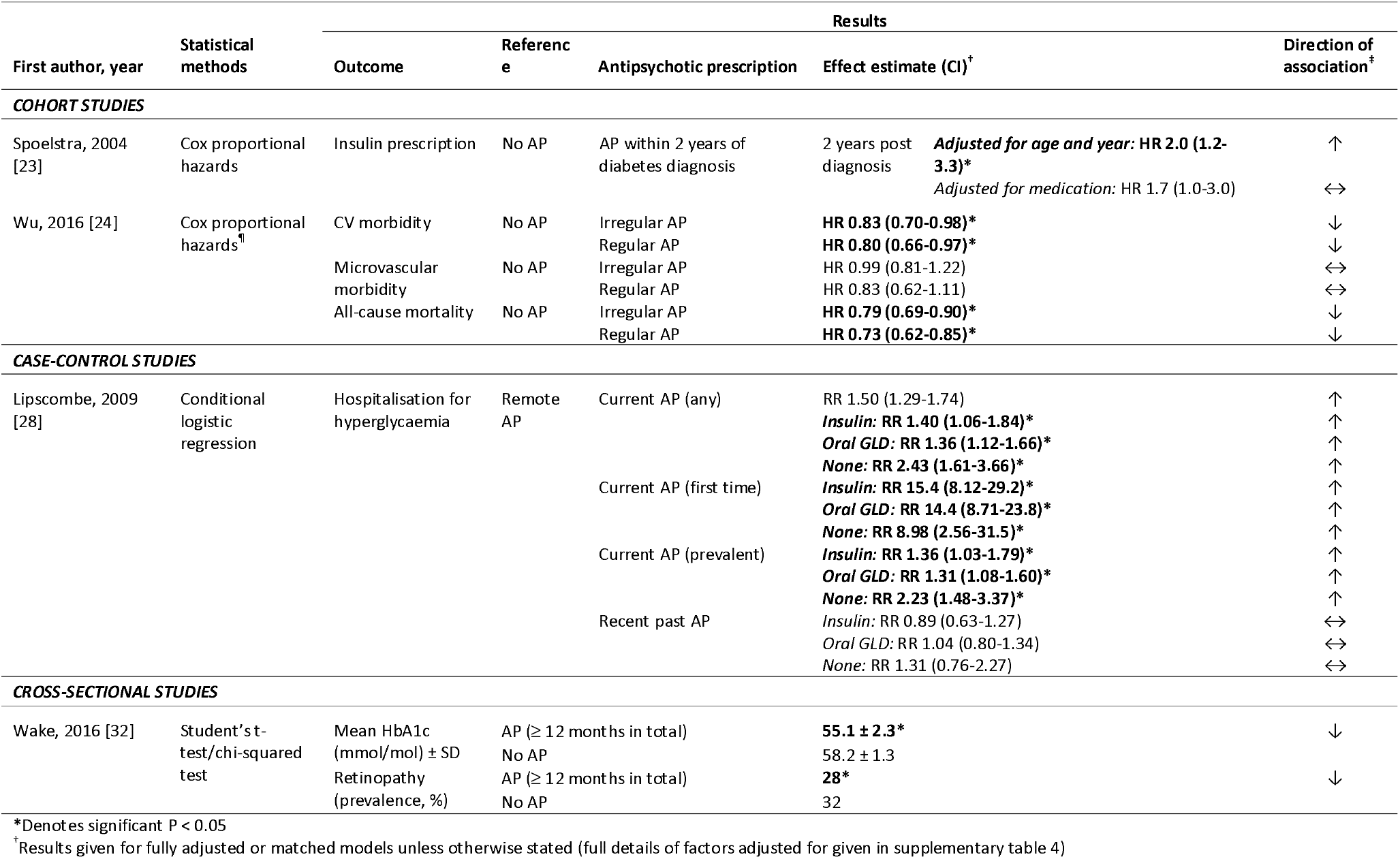
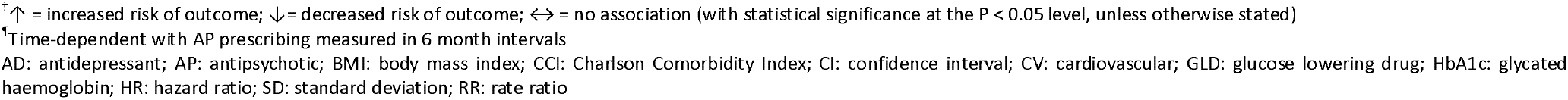
Results of observational studies reporting on the association between antipsychotic drug prescribing and outcomes in people with diabetes

A second, cross-sectional, study found that the prevalence of retinopathy was lower in people who had been prescribed antipsychotics for a total of more than 12 months than in a comparison group matched for age, sex, diabetes type and duration, body mass index and smoking status that had not been prescribed antipsychotics (Table 3) [32].

Two studies reported a link between that antipsychotic prescribing and poorer glycaemic control [23, 28], and one reported the opposite finding [32]. The only cohort study [23] found that receipt of antipsychotic prescriptions within the first two years of diabetes diagnosis was associated with a two-fold increased risk of first insulin prescription, which attenuated to a 30% increased risk after adjustment for other drug prescriptions (Table 3). In a case-control study of older adults, receipt of an antipsychotic prescription in the previous 180 days was associated with an increased risk of hospitalisation for hyperglycaemia, regardless of diabetes medication type [28]. This effect was strongest among those receiving a first prescription of antipsychotics (Table 3) and consistent across antipsychotic subtypes (Supplemental Table 4b). In contrast, a cross-sectional study found that mean HbA1c values were significantly lower in people prescribed antipsychotics for more than 12 months in total than in those who never received antipsychotics [32].

#### 3.4.3 Other cardiovascular risk factors

Antipsychotic prescribing was also associated with significantly lower systolic and diastolic blood pressure and total cholesterol although absolute differences were small (Supplemental Table 5b).

## 4.1 DISCUSSION

Our review revealed that few observational studies have investigated the association between antidepressant and antipsychotic drug prescribing and diabetes outcomes. Studies of associations with microvascular complications and cardiometabolic factors other than glycaemic control and the association between antipsychotic prescribing and any diabetes complication were particularly limited. The published studies are highly heterogeneous, particularly in terms of the study population, exposure definitions, and outcomes. Lack of comparability and study shortcomings highlight key gaps and limit conclusions. Tentative conclusions are that, among people with diabetes, antidepressant prescribing may be associated with an increased risk of cardiovascular morbidity or mortality, and both antidepressant and antipsychotic prescribing may be linked to poorer glycaemic control, but the evidence is very mixed.

### 4.1 Strengths and limitations

To the best of our knowledge this is the first systematic review of observational studies reporting associations between antidepressant or antipsychotic drug prescribing and complications, mortality, or cardiometabolic risk factors in people with diabetes. Other strengths include our use of a detailed and comprehensive search strategy applied to three bibliographic databases, independent screening and data extraction by two reviewers, and assessment of study quality.

A limitation of this review is the exclusion of outcomes related to weight changes and obesity, that have previously been linked to antidepressant and antipsychotic use [34, 35], as these outcomes were beyond the scope of our review. Also beyond our scope were studies which reported on progression of diabetes complications or compared prescribing of one psychotropic drug versus another. The latter is particularly important when investigating adverse effects of antidepressant and antipsychotic prescribing in people with SMI, where given the clinical need to prescribe, comparison of type of medication becomes more important than comparison of use versus non-use. We recommend that future reviews examine this topic in greater depth by focusing on study populations with comorbid SMI and diabetes only.

Further limitations of this review reflect the limitations of existing studies. Many of these have insufficient control of confounding factors, including lifestyle behaviours, socioeconomic status, and critically, confounding by indication, particularly for mental illness. It is difficult to estimate how the latter might have affected estimates with potential bias in both directions. Mental illness is associated with higher risk of poor diabetes outcomes [36-38], partly through higher prevalence of unhealthy lifestyle factors, including overweight/obesity, lower socioeconomic status [39], poorer treatment adherence, and in some settings, receipt of sub-optimal care [39]. Conversely, in other settings, people with comorbid mental illness and diabetes may be more likely than people with diabetes without mental illness to receive optimal routine monitoring of certain care indicators [40], perhaps due to more frequent visits to their primary care practitioner [41].

Although ascertainment of prescriptions through electronic records minimises information bias and loss to follow-up, prescribing records do not reflect actual drug use. For studies where exposure status was identified using interviews or assessment visits, this information may be subject to social desirability and recall biases. Misclassifying people as exposed, either where people do not take drugs as prescribed or fail to disclose the drugs prescribed, would weaken effect estimates. Similarly, electronic records often only include outcomes resulting in hospitalisation and may also be subject to misclassification bias.

Some studies had low precision due to small sample sizes, which affects interpretation of effect estimates, especially for less common microvascular complications, as well as for the investigation of differences by drug subtypes.

Finally, in anticipation of limited evidence, we included cross-sectional studies in our review. It is however possible that diabetes complications increase the risk of a person developing mental illness, such as depression [36], and in turn the likelihood of being prescribed antidepressant or antipsychotic medications.

### 4.2 Interpretation of findings: antidepressants

Our findings on antidepressant use and reduced risk of cardiovascular morbidity or mortality in people with diabetes contrasts with previous reviews which report the opposite in general populations that included people with and without diabetes [9, 42]. These contrasting findings may reflect key differences in associations for different population sub-groups. Indeed, pooled results of only studies including people with, or at high risk of, cardiovascular disease similarly found that antidepressant use/prescribing was associated with a statistically non-significant reduced risk of cardiovascular morbidity [42]. Systematic reviews of trials suggest that SSRI use can improve short-term glycaemic control in people with diabetes [13, 14]. Some of the observational studies we identified found similar results but others found no differences in glycaemic control between groups, perhaps related to study population, design, sample size and approaches to control of confounding. 4.3 Interpretation of findings: antipsychotics

Our review revealed the sparse evidence on antipsychotic prescribing and diabetes outcomes. Although based on just one cohort study [24], the association between antipsychotic prescribing and lower mortality risk in people with schizophrenia and diabetes aligns with findings on antipsychotic use and lower long-term mortality in people with schizophrenia [43, 44] and short-term mortality in the general population [45]. However, in contrast to previous reports of a link between antipsychotic use and increased risk of cardiovascular disease in general populations [10, 46], this cohort study found the opposite. Although counter-intuitive, given their adverse metabolic and cardiovascular effects [8, 34, 47], antipsychotic use alongside psychological support in people with SMI may improve physical and psychosocial functioning, thereby improving lifestyle and compliance with diabetes care and treatment, which in turn could reduce risk of diabetes complications.. The few studies on antipsychotic prescribing suggest an association with increased risk of poorer glycaemic control, which aligns with established adverse glycaemic effects of some antipsychotics [48].

### 4.3 Implications

The sparse existing literature on the association between antidepressant and antipsychotic prescribing and diabetes outcomes limits our conclusions and implications for practice. There are major gaps that should be addressed in future research in order to inform clinical practice. The increased use of antidepressant and antipsychotic medications in the general population is concerning given the evident lack of understanding of potential for adverse effects and increasing prevalence of diabetes. The striking gap in the evidence on antipsychotic drug prescribing in relation to diabetes outcomes in particular should be urgently addressed. Antidepressant and antipsychotic drugs are an essential component of the treatment of mental health conditions, and timely treatment in people with diabetes is crucial, given the poor diabetes outcomes for people with both diabetes and mental ill health [49]. It is therefore important to establish treatment-associated risks of diabetes outcomes within population sub-groups to inform and enhance drug prescribing and monitoring practices for diabetes and its complications. To improve upon existing studies and advance our understanding, future studies should: be well-powered (particularly to investigate differences); include information on mental illness, lifestyle, and other confounding factors; distinguish between drug subtypes; assess whether and how risk changes over time; and consider the potential effects of cumulative exposures.

### 4.5 Conclusions

Few studies have described the association between antidepressant and antipsychotic medications and diabetes outcomes, with shortcomings and mixed findings limiting the ability to draw conclusions. While future research addresses this evidence gap, clinicians, including diabetologists, primary care practitioners, and where appropriate, psychiatrists, should attempt to ensure regular monitoring of cardiometabolic risk factors and early indicators of complications in people with diabetes who are being treated with antidepressant or antipsychotic medication (irrespective of indication).

## Data Availability

All data produced in the present work are contained in the manuscript

