## Supplementary material for "Antidepressant and antipsychotic drug prescribing and complications of diabetes: a systematic review of observational studies"

*Corresponding author:

Supplementary Material

**Supplemental appendix 1** Electronic search strategies

**Supplemental Table 1** Study inclusion and exclusion criteria (PICO framework)

**Supplemental Table 2** (a): Antidepressant drugs included in relevant studies

### Supplemental Table 2 (b): Antipsychotic drugs included in relevant studies

### Supplemental Table 3: Assessment of study quality: Newcastle-Ottawa Scale summary

### Supplemental Table 4: Additional information on adjustment for confounding in statistical models

**Supplementary Table 5** (a): Supplementary results of observational studies reporting on the association between antidepressant drug prescribing and outcomes in people with diabetes

### Supplemental Table 5 (b): Supplementary results of observational studies reporting on the association between antipsychotic drug prescribing and outcomes in people with diabetes

**Supplemental appendix 1** Electronic search strategies

**PubMed Search Strategy**

“diabetes mellitus”[MeSH Terms] OR “diabetes”[All Fields] OR “diabetic”[All Fields]

“antidepressive agents”[Mesh] OR “antidepressants”[All Fields] OR “anti-depressants”[All Fields] OR “anti depressants”[All Fields] OR “antidepressant”[All Fields] OR “anti-depressant”[All Fields] OR “anti depressant”[All Fields] OR "antidepressive" OR "anti-depressive" OR "anti depressive"

“serotonin uptake inhibitors”[Pharmacological Action] OR “serotonin uptake inhibitors”[MeSH Terms] OR “selective serotonin reuptake inhibitors”[All Fields] OR “selective serotonin reuptake inhibitor”[All Fields] OR “SSRI”[All Fields]

“serotonin and noradrenaline reuptake inhibitors”[Pharmacological Action] OR “serotonin and noradrenaline reuptake inhibitors”[MeSH Terms] OR “serotonin and noradrenaline reuptake inhibitors”[All Fields] OR “serotonin and noradrenaline reuptake inhibitor”[All Fields] OR “SNRI”[All Fields]

“monoamine oxidase inhibitors”[Pharmacological Action] OR “monoamine oxidase inhibitors”[MeSH Terms] OR “monoamine oxidase inhibitors”[All Fields] OR “monoamine oxidase inhibitor”[All Fields] OR “MAOI”[All Fields]

(“tricyclic” OR “tetracyclic”) AND (“antidepressants”[All Fields] OR “anti-depressants”[All Fields] OR “anti depressants”[All Fields] OR “antidepressant”[All Fields] OR “anti-depressant”[All Fields] OR “anti depressant”[All Fields])

“agomelatine”[All Fields] OR “valdoxan”[All Fields]

“amitriptyline”[MeSH Terms] OR “amitriptyline”[All Fields] OR “triptafen”[All Fields]

“amoxapine”[MeSH Terms] OR “amoxapine”[All Fields] OR “asendin”[All Fields]

“bupropion”[MeSH Terms] OR “bupropion”[All Fields]

“citalopram”[MeSH Terms] OR “citalopram”[All Fields]

“clomipramine”[MeSH Terms] OR “clomipramine”[All Fields] OR “anafranil”[All Fields]

"desipramine"[MeSH Terms] OR "desipramine"[All Fields] OR “norpramin”[All Fields]

“dothiepin”[MeSH Terms] OR “dothiepin”[All Fields] OR “dosulepin”[All Fields] OR “prothiaden”[All Fields]

“doxepin”[MeSH Terms] OR “doxepin”[All Fields]

“duloxetine hydrochloride”[MeSH Terms] OR “duloxetine”[All Fields] OR “cymbalta”[All Fields]

“escitalopram”[All Fields] OR “cipralex”[All Fields]

“fluoxetine”[MeSH Terms] OR “fluoxetine”[All Fields] OR “prozac”[All Fields]

“flupenthixol”[MeSH Terms] OR “flupenthixol”[All Fields] OR “fluanxol”[All Fields]

“fluvoxamine”[MeSH Terms] OR “fluvoxamine”[All Fields] OR “faverin”[All Fields]

“imipramine”[MeSH Terms] OR “imipramine”[All Fields]

“isocarboxazid”[MeSH Terms] OR “isocarboxazid”[All Fields]

“lofepramine”[MeSH Terms] OR “lofepramine”[All Fields]

“mianserin”[MeSH Terms] OR “mianserin”[All Fields] OR “tolvon”[All Fields]

“mirtazapine”[All Fields] OR “remeron”[All Fields] OR “zispin”[All Fields]

“moclobemide”[MeSH Terms] OR “moclobemide”[All Fields] OR “manerix”[All Fields]

“nortriptyline”[MeSH Terms] OR “nortriptyline”[All Fields] OR “allegron”[All Fields]

“paroxetine”[MeSH Terms] OR “paroxetine”[All Fields] OR “seroxat”[All Fields]

“phenelzine”[MeSH Terms] OR “phenelzine”[All Fields] OR “nardil”[All Fields]

“reboxetine”[Supplementary Concept] OR “reboxetine”[All Fields]

“sertraline”[MeSH Terms] OR “sertraline”[All Fields] OR “lustral”[All Fields]

“tranylcypromine”[MeSH Terms] OR “tranylcypromine”[All Fields]

“trazodone”[MeSH Terms] OR “trazodone”[All Fields] OR “molipaxin”[All Fields]

“trimipramine”[MeSH Terms] OR “trimipramine”[All Fields] OR “surmontil”[All Fields]

“venlafaxine hydrochloride”[MeSH Terms] OR “venlafaxine”[All Fields] OR “effexor”[All Fields]

“vortioxetine”[All Fields] OR “trintellix”[All Fields] OR “brintellix”[All Fields]

2 OR 3 OR 4 OR 5 OR 6 OR 7 OR 8 OR 9 OR 10 OR 11 OR 12 OR 13 OR 14 OR 15 OR 16 OR 17 OR 18 OR 19 OR 20 OR 21 OR 22 OR 23 OR 24 OR 25 OR 26 OR 27 OR 28 OR 29 OR 30 OR 31 OR 32 OR 33 OR 34 OR 35 OR 36

“antipsychotic agents”[Mesh] OR “antipsychotics”[All Fields] OR “anti-psychotics”[All Fields] OR “anti psychotics”[All Fields] OR “antipsychotic”[All Fields] OR “anti-psychotic”[All Fields] OR “anti psychotic”[All Fields] OR “neuroleptics”[All Fields] OR “neuroleptic”[All Fields] OR “major tranquilizers”[All Fields] OR “major tranquilizer”[All Fields]

“amisulpride”[All Fields] OR “solian”[All Fields]

“aripiprazole”[MeSH Terms] OR “aripiprazole”[All Fields] OR “abilify”[All Fields]

“asenapine”[All Fields] OR “saphris”[All Fields] OR “sycrest”[All Fields]

“benperidol”[MeSH Terms] OR “benperidol”[All Fields] OR “anquil”[All Fields]

“chlorpromazine”[MeSH Terms] OR “chlorpromazine”[All Fields] OR “largactil”[All Fields]

“clopenthixol”[MeSH Terms] OR “clopenthixol”[All Fields] OR “clopentixol”[All Fields] OR “sordinol”[All Fields] OR “ciatyl”[All Fields]

“clozapine”[MeSH Terms] OR “clozapine”[All Fields] OR “clozaril”[All Fields]

“flupenthixol”[MeSH Terms] OR “flupenthixol”[All Fields] OR “flupentixol”[All Fields] OR “fluanxol”[All Fields] OR “depixol”[All Fields]

“haloperidol”[MeSH Terms] OR “haloperidol”[All Fields] OR “haldol”[All Fields]

“lurasidone hydrochloride”[MeSH Terms] OR “lurasidone”[All Fields] OR “latuda”[All Fields]

“methotrimeprazine”[MeSH Terms] OR “methotrimeprazine”[All Fields] OR “levomepromazine”[All Fields]

“olanzapine”[All Fields] OR “zalasta”[All Fields] OR “zyprexa”[All Fields]

“paliperidone palmitate”[MeSH Terms] OR “paliperidone” OR “trevicta”[All Fields]

“periciazine”[All Fields] OR “pericyazine”[All Fields]

“perphenazine”[MeSH Terms] OR “perphenazine”[All Fields]

“pimozide”[MeSH Terms] OR “pimozide”[All Fields] OR “orap”[All Fields]

“prochlorperazine”[MeSH Terms] OR “prochlorperazine”[All Fields] OR “buccastem”[All Fields] OR “stemetil”[All Fields]

“promazine”[MeSH Terms] OR “promazine”[All Fields] OR “sparine”[All Fields]

“quetiapine fumarate”[MeSH Terms] OR “quetiapine”[All Fields] OR “seroquel”[All Fields]

“risperidone”[MeSH Terms] OR “risperidone”[All Fields] OR “risperdal”[All Fields]

“sulpiride”[All Fields] OR “dogmatil”[All Fields] OR “dolmatil”[All Fields] OR “sulpor”[All Fields]

“trifluoperazine”[MeSH Terms] OR “trifluoperazine”[All Fields] OR “stelazine”[All Fields]

“ziprasidone”[All Fields] OR “geodon”[All Fields]

“zuclopenthixol “[All Fields] OR “clopixol”[All Fields] OR “cisordinol”[All Fields] OR “acuphase”[All Fields]

38 OR 39 OR 40 OR 41 OR 42 OR 43 OR 44 OR 45 OR 46 OR 47 OR 48 OR 49 OR 50 OR 51 OR 52 OR 53 OR 54 OR 55 OR 56 OR 57 OR 58 OR 59 OR 60 OR 61 OR 62

37 OR 63

1 AND 64

**EMBASE Search Strategy**

exp diabetes mellitus/

diabet*.tw.

or/1-2

antidepressant agent/

(antidepress* or anti-depress* or anti depress*).tw.

exp serotonin uptake inhibitor/

exp noradrenalin uptake inhibitor/

exp monoamine oxidase inhibitor/

exp tricyclic antidepressant agent/

exp tetracyclic antidepressant agent/

(agomelatine or valdoxan).tw.

(amitriptyline or triptafen).tw.

(amoxapine or asendin).tw.

bupropion.tw.

citalopram.tw.

(clomipramine or anafranil).tw.

(desipramine or norpramin).tw.

(dothiepin or dosulepin or prothiaden).tw.

doxepin.tw.

(duloxetine hydrochloride or cymbalta).tw.

(escitalopram or cipralex).tw.

(fluoxetine or prozac).tw.

(flupenthixol or fluanxol).tw.

(fluvoxamine or faverin).tw.

imipramine.tw.

isocarboxazid.tw.

lofepramine.tw.

(mianserin or tolvon).tw.

(mirtazapine or remeron or zispin).tw.

(moclobemide or manerix).tw.

(nortriptyline or allegron).tw.

(paroxetine or seroxat).tw.

(phenelzine or nardil).tw.

reboxetine.tw.

(sertraline or lustral).tw.

tranylcypromine.tw.

(trazodone or molipaxin).tw.

(trimipramine or surmontil).tw.

(venlafaxine or effexor).tw.

(vortioxetine or trintellix or brintellix).tw.

antipsychotic agent/

(antipsychotic* or anti-psychotic* or anti psychotic* or neuroleptic* or major tranquilizer*).tw.

(amisulpride or solian).tw.

(aripiprazole or abilify).tw.

(asenapine or saphris or sycrest).tw.

(benperidol or anquil).tw.

(clopenthixol or clopentixol or sordinol or ciatyl).tw.

(clozapine or clozaril).tw.

(flupenthixol or flupentixol or fluanxol or depixol).tw.

(haloperidol or haldol).tw.

(lurasidone or latuda).tw.

(methotrimeprazine or levomepromazine).tw.

(olanzapine or zalasta or zyprexa).tw.

(paliperidone or trevicta).mp

(periciazine or pericyazine).tw.

perphenazine.tw.

(pimozide or orap).tw.

(prochlorperazine or buccastem or stemetil).tw.

(promazine or sparine).tw.

(quetiapine or seroquel).tw.

(risperidone or risperdal).tw.

(sulpiride or dogmatil or dolmatil or sulpor).tw.

(trifluoperazine or stelazine).tw.

(ziprasidone or geodon).tw.

(zuclopenthixol or clopixol or cisordinol or acuphase).tw.

or/4-65

outcome assessment/

health status/

mortality/

hospital admission/

haemoglobin A1c/

diabetic control/

blood pressure/

total cholesterol level/

triacylglycerol level/

ischemic heart disease/

cerebrovascular accident/

peripheral occlusive artery disease/

exp diabetic complication/

outcome*.tw.

mortality.tw.

hospital admission.tw.

(blood glucose or blood sugar).tw.

blood pressure.tw.

cholesterol.tw.

triglyceride adj2 levels.tw.

(myocardial infarction or heart attack).tw.

stroke.tw.

peripheral adj3 disease.tw.

diabetic foot.tw.

(diabet* ketoacidosis or DKA).tw.

retinopathy.tw.

neuropathy.tw.

nephropathy.tw.

or/67-94

3 and 66 and 96

**PsycInfo Search Strategy**

exp diabetes mellitus/

diabet*.tw.

or/1-2

exp antidepressant drugs/

(antidepress* or anti-depress* or anti depress*).tw.

(agomelatine or valdoxan).tw.

(amitriptyline or triptafen).tw.

(amoxapine or asendin).tw.

bupropion.tw.

citalopram.tw.

(clomipramine or anafranil).tw.

(desipramine or norpramin).tw.

(dothiepin or dosulepin or prothiaden).tw.

doxepin.tw.

(duloxetine hydrochloride or cymbalta).tw.

(escitalopram or cipralex).tw.

(fluoxetine or prozac).tw.

(flupenthixol or fluanxol).tw.

(fluvoxamine or faverin).tw.

imipramine.tw.

isocarboxazid.tw.

lofepramine.tw.

(mianserin or tolvon).tw.

(mirtazapine or remeron or zispin).tw.

(moclobemide or manerix).tw.

(nortriptyline or allegron).tw.

(paroxetine or seroxat).tw.

(phenelzine or nardil).tw.

reboxetine.tw.

(sertraline or lustral).tw.

tranylcypromine.tw.

(trazodone or molipaxin).tw.

(trimipramine or surmontil).tw.

(venlafaxine or effexor).tw.

(vortioxetine or trintellix or brintellix).tw.

exp antipsychotic drugs/

(antipsychotic* or anti-psychotic* or anti psychotic* or neuroleptic* or major tranquilizer*).tw.

(amisulpride or solian).tw.

(aripiprazole or abilify).tw.

(asenapine or saphris or sycrest).tw.

(benperidol or anquil).tw.

(clopenthixol or clopentixol or sordinol or ciatyl).tw.

(clozapine or clozaril).tw.

(flupenthixol or flupentixol or fluanxol or depixol).tw.

(haloperidol or haldol).tw.

(lurasidone or latuda).tw.

(methotrimeprazine or levomepromazine).tw.

(olanzapine or zalasta or zyprexa).tw.

(paliperidone or trevicta).mp

(periciazine or pericyazine).tw.

perphenazine.tw.

(pimozide or orap).tw.

(prochlorperazine or buccastem or stemetil).tw.

(promazine or sparine).tw.

(quetiapine or seroquel).tw.

(risperidone or risperdal).tw.

(sulpiride or dogmatil or dolmatil or sulpor).tw.

(trifluoperazine or stelazine).tw.

(ziprasidone or geodon).tw.

(zuclopenthixol or clopixol or cisordinol or acuphase).tw.

or/4-60

mortality/

hospital admission/

blood sugar/

exp blood pressure/

total cholesterol level/

lipids/

cerebrovascular accidents/

exp cardiovascular disorders/

outcome*.tw.

mortality.tw.

hospital admission.tw.

(blood glucose or blood sugar).tw.

blood pressure.tw.

cholesterol.tw.

triglyceride adj2 levels.tw.

(myocardial infarction or heart attack).tw.

stroke.tw.

peripheral adj3 disease.tw.

diabetic foot.tw.

(diabet* ketoacidosis or DKA).tw.

retinopathy.tw.

neuropathy.tw.

nephropathy.tw.

or/62-84

3 and 61 and 85

**Supplemental Table 1** Study inclusion and exclusion criteria (PICO framework)

|  | **Inclusion** | | **Exclusion** |
| --- | --- | --- | --- |
| **Population** | Adults with pre-existing type 2 diabetes mellitus in any location or setting | | Children (< 18 years); people with type 1 diabetes mellitus; studies that did not report separately on outcomes in people with diabetes; studies where the study population was selected on the presence of a specific disease of interest and where data on psychotropic medication prescribing was reported for a sub-set of the study population with diabetes but without the provision of key descriptive characteristics of the sub-set. |
| **Intervention** | All type antidepressant and/or antipsychotic drug use or prescribing | | Lithium use or prescribing only |
| **Comparison** | The absence of antidepressant and/or antipsychotic drug use or prescribing | | A second antidepressant and/or antipsychotic drug; no comparison group |
| **Outcome** | *Clinical complications*  *(primary)* | Cardiovascular morbidity; retinopathy; neuropathy; nephropathy; all-cause mortality; cause-specific mortality | Outcomes related to the progression of clinical complications; weight changes/obesity; outcomes related to mental wellbeing |
|  | *Cardiometabolic risk factors (secondary)* | Glycaemic control (HbA1c, hyperglycaemia, insulin initiation); blood pressure (hypertension, systolic/diastolic); lipid levels (dyslipidaemia, low-density/high-density lipoprotein cholesterol, total cholesterol, triglycerides) |  |
| Study design | Observational studies (cohort, case-control, cross-sectional); accessible in the English language | | Case reports; experimental studies; secondary studies (narrative reviews, literature reviews, meta-analyses) |

**Supplemental Table 2** (a): Details of antidepressant drugs included in studies reporting on antidepressant prescribing in relation to diabetes outcomes

| First author, publication year, study setting  [study period] | Method | SSRIs (ATC N06A-) | TCAs (ATC N06A-) | Other (ATC N06A-) |
| --- | --- | --- | --- | --- |
| Higgins, 2007 [27] USA  [July 2002] | NR | NR | NR | NR |
| Rubin, 2010, 2013 [21] USA  [June 2001 to approx. 2005] | NR | NR | NR | NR |
| Noordam, 2016* [31] Netherlands  [1991 to 2012] | ATC | Paroxetine (B05) | Amitriptyline (A09) | NR |
| Rådholm, 2015 [19] Sweden  [January 2008 to December 2010] | ATC | NR | NR | NR |
| Yekta, 2015 [30] USA  [2005 to 2008] | NR | Fluoxetine (B03) Citalopram (B04) Paroxetine (B05) Sertraline (B06) Fluvoxamine (B08) Escitalopram (B10) | Desipramine (A01) Imipramine (A02) Clomipramine (A04) Trimipramine (A06) Amitriptyline (A09) Nortriptyline (A10) Protriptyline (A11) Doxepin (A12) Maprotiline (A21) | Trazodone (X05) Nefazodone (X06) Mirtazapine (X11) Bupropion (X12) Venlafaxine (X16) Duloxetine (X21) |
| Brieler, 2016 [15] USA  [July 2008 to July 2013] | NR | Fluoxetine (B03) Citalopram (B04) Paroxetine (B05) Sertraline (B06) Fluvoxamine (B08) Escitalopram (B10) | Desipramine (A01) Imipramine (A02) Clomipramine (A04) Amitriptyline (A09) Nortriptyline (A10) Doxepin (A12) | Trazodone (X05) Nefazodone (X06) Mirtazapine (X11) Bupropion (X12) Venlafaxine (X16) Duloxetine (X21) Desvenlafaxine (N06AX23) |
| Kammer, 2016* [28] USA  [2002 to 2009] | NR | Fluoxetine (B03) Paroxetine (B05) Sertraline (B06) | NR | NR |
| Würtz, 2016 [25] Denmark  [July 2004 to December 2012] | ATC | Zimeldine (B02) Fluoxetine (B03) Citalopram (B04) Paroxetine (B05) Sertraline (B06) Alaproclate (B07) Fluvoxamine (B08) Etoperidone (B09) Escitalopram (B10) | None | None |
| Hazuda, 2019 [18]  USA  [June 2001 to September 2012] | NR | NR | NR | NR |
| Rohde, 2021 [20] Denmark [January 2000 to October 2016] | ATC | Fluoxetine (B03) Citalopram (B04) Paroxetine (B05) Sertraline (B06) Fluvoxamine (B08) Escitalopram (B10) | Imipramine (A02) Clomipramine (A04) Amitriptyline (A09) Nortriptyline (A10) | Mianserin (X03) Mirtazapine (X11) Venlafaxine (X16) Reboxetine (X18) Duloxetine (X21) |
| Chen, 2021 [16]  Taiwan  [January 1997 to December 2010] | ATC | NR | NR | NR |
| Wu, 2021 [24]  Taiwan  [2001 to 2004] | ATC | Fluoxetine (B03) Citalopram (B04) Paroxetine (B05) Sertraline (B06) Fluvoxamine (B08) | Imipramine (A02) Clomipramine (A04) Amitriptyline (A09) Doxepin (A12) Dothiepin (A16) Melitracen (A14) Maprotiline (A21) | Trazodone (X05) Nefazodone (X06) Mirtazapine (X11) Bupropion (X12) Venlafaxine (X16) Milnacipran (X17) Duloxetine (X21) Moclobemide (G02) |
| Chen, 2022 [17]  Taiwan  [1999 to 2013] | NR | NR | NR | NR |
| Rohde, 2022 [26]  Denmark  [January 2000 to October 2016] | ATC | Fluoxetine (B03) Citalopram (B04) Paroxetine (B05) Sertraline (B06) Fluvoxamine (B08) Escitalopram (B10) | Imipramine (A02) Clomipramine (A04) Amitriptyline (A09) Nortriptyline (A10) | Mianserin (X03) Mirtazapine (X11) Venlafaxine (X16) Reboxetine (X18) Duloxetine (X21) |

*List not comprehensive, authors specified commonly prescribed drugs only

AD: antidepressant; ATC: Anatomical Therapeutic Chemical; MAOI: monoamine oxidase inhibitor; NR: not reported; SNRI: serotonin–norepinephrine reuptake inhibitor; SSRI: selective serotonin reuptake inhibitor; TCA: tricyclic antidepressant

NB. The ATC classification system is the gold standard for identifying drugs in international drug research. It is maintained by the World Health Organization Collaborating Centre for Drug Statistics Methodology ([www.whocc.no](http://www.whocc.no)). ADs are classified at the third level, N06A, and divided into groups at higher levels according to their therapeutic, pharmacological, and chemical properties. Relevant groups include SSRIs (N06AB-), TCAs (N06AA-), MAOIs, (N06AF/G-), and other (N06AX-). Drugs including venlafaxine (X16) and duloxetine (X21) may also be classified as SNRIs.

### Supplemental Table 2 (b): Details of antipsychotic drugs included in studies reporting on antipsychotic prescribing in relation to diabetes outcomes

| First author, publication year, study setting  [study period] | Method of identification | FGAs (ATC N05A-) | SGAs (ATC N05A-) |
| --- | --- | --- | --- |
| Spoelstra, 2004 [22]  Netherlands  [January 1991 to June 1999] | ATC | NR | Clozapine (H02) Olanzapine (H03) Quetiapine (H04) Risperidone (X08) |
| Lipscombe, 2009* [32]  Canada  [April 2002 to March 2006] | NR | NR | Olanzapine (H03) Quetiapine (H04) Risperidone (X08) |
| Wake, 2016 [29]  UK  [2010] | BNF subsection 4.2.^†‡^ | NR | NR |
| Wu, 2016 [23]  Taiwan  [2001 to 2012] | ATC | Chlorpromazine (A01) Levomepromazine (A02) Promazine (A03) Fluphenazine (B02) Perphenazine (B03) Prochlorperazine (B04) Trifluoperazine (B06) Thioridazine (C02) Pipotiazine (C04) Haloperidol (D01) Flupentixol (F01) Zuclopenthixol (F05) Pimozide (G02) Loxapine (H01) | Clozapine (H02) Olanzapine (H03) Quetiapine (H04) Sulpiride (L01) Amisulpride (L05) Risperidone (X08) Zotepine (X11) |

*Included APs funded by the Ontario Drug Benefit programme during the study period

^†^The BNF is a reference for drug prescribing used in the UK

^‡^Subsection 4.2.1 refers to APs, this excludes: depot injection formulations (BNF subsection 4.2.2); prochlorperazine (B04) and droperidol (D08) (BNF section 4.6, drugs used for vertigo); and asenapine (H05) and lithium (N01) (BNF section 4.2.3, drugs used for mania and hypomania)

AP: antipsychotic; ATC: Anatomical Therapeutic Chemical; BNF: British National Formulary; FGA: first-generation antipsychotic; NR: not reported; SGA: second-generation antipsychotic

NB. The ATC classification system is the gold standard for identifying drugs in international drug research. It is maintained by the World Health Organization Collaborating Centre for Drug Statistics Methodology ([www.whocc.no](http://www.whocc.no)). APs are classified at the third level, N05A, and divided into groups at higher levels according to their therapeutic, pharmacological, and chemical properties. Relevant groups include FGAs and SGAs.

### Supplemental Table 3: Assessment of study quality: Newcastle-Ottawa Scale summary

| First author, year | Selection | Comparability | Outcome/Exposure | Total stars |
| --- | --- | --- | --- | --- |
| *COHORT STUDIES* | | | | |
| Spoelstra, 2004 [22] | **** | * | *** | 8/9 |
| Rubin, 2010, 2013 [21] | *** | * | ** | 6/9 |
| Rådholm, 2015 [19] | **** |  | *** | 7/9 |
| Brieler, 2016 [15] | **** | * | *** | 8/9 |
| Wu, 2016 [23] | **** | * | *** | 8/9 |
| Würtz, 2016 [25] | **** |  | *** | 7/9 |
| Hazuda, 2019 [18] | *** | * | ** | 6/9 |
| Rohde, 2021 [20] | **** | * | *** | 8/9 |
| Chen, 2021 [16] | **** | * | *** | 8/9 |
| Wu, 2021 [24] | **** | * | ** | 7/9 |
| Chen, 2022 [17] | **** | * | *** | 8/9 |
| Rohde, 2022 [26] | **** | * | ** | 7/9 |
| *CASE-CONTROL STUDIES* | | | | |
| Lipscombe, 2009 [32] | **** | * | *** | 8/9 |
| Noordam, 2016 [31] | **** | * | *** | 8/9 |
| *CROSS-SECTIONAL STUDIES* | | | | |
| Higgins, 2007 [27] | *** |  | ** | 5/9 |
| Yekta, 2015 [30] | ** | * | *** | 6/9 |
| Kammer, 2016 [28] |  | * | ** | 3/9 |
| Wake, 2016 [29] | **** | * | * | 6/9 |

Studies were awarded a maximum of four stars for study selection, two stars for group comparability, and three stars for ascertainment of outcome or exposure, to a maximum total of nine. Separate versions of the scale were used for cohort, case-control, and cross-sectional studies.

### Supplemental Table 4 (a) Additional information on adjustment for confounding in statistical models for the association between antidepressant prescribing and outcomes

| First author, year | Statistical methods | Confounders adjusted for in analyses | Details of composite outcomes (where relevant) |
| --- | --- | --- | --- |
| *COHORT STUDIES* | | | |
| Rubin, 2010, 2013 [22] | GEE | Age; sex; ethnicity; education; HbA1c (prior year); history of CVD; duration of diabetes; AD prescription (prior year); year of follow-up | NA |
| Rådholm, 2015 [20] | Descriptive | Stratified by age and sex | NA |
| Brieler, 2016 [16] | GEE | Age; sex; ethnicity; anxiety disorder; obesity; hyperlipidaemia; hypertension; vascular disease; referral to dietary education; smoking; insulin prescription; other GLD prescription; primary care clinic utilisation | NA |
| Wu, 2016 [24] |  |  | Cardiovascular morbidity (hospitalisation for coronary heart disease and stroke; peripheral vascular disease with stent insertion; vascular shunt or bypass; vessel repair procedure), microvascular morbidity (retinopathy; blindness; end-stage renal disease with dialysis; vessel operations for haemodialysis; kidney transplantation; hospitalisation for diabetic foot infection; lower extremity amputations), all-cause mortality |
| Würtz, 2016 [26] | Cox proportional hazards | Previous MI, atrial ﬁbrillation or ﬂutter; intermittent arterial claudication; dementia; CCI (excluding MI, PVD, and dementia); other drug prescriptions | NA |
| Hazuda, 2019 [19] | Cox proportional hazards | Age; ethnicity; history of CVD; HbA1c; BMI; waist circumference; insulin prescription; hypercholesterolaemia; hypertension; smoking; estimated exercise stress level; HRT prescription (women only); clinic attended; assigned clinical trial intervention | xxx |
| Chen, 2021 [17] | Cox proportional hazards | Age; sex; income; urbanization; hypertension; hypercholesterolaemia; CAD; CKD; heart failure; peptic ulcer; aspirin; plavix | NA |
| Rohde, 2021 [21] | Cox proportional hazards | Age; sex; marital status; HbA1c (baseline); LDL levels (baseline); obesity; kidney functioning; CCI (excluding diabetes); diabetes complications; alcohol-related disorders; smoking-associated disorders; other drug prescriptions | NA |
| Wu, 2021 [25] | None | Crude incidence in comparison groups | Macrovascular complications (ischaemic heart disease, stroke, peripheral vascular disease, vascular shunts or bypasses, vessel repair procedures), microvascular complications (diabetic retinopathies, end-stage renal disease, diabetes foot infections), all-cause mortality |
| Chen 2022 [18] | Cox proportional hazards | Age; sex; economic level; urbanization level; CCI score; hypertension; acute MI, hyperlipidemia; atrial fibrillation; COPD; depression; bipolar disorder; schizophrenia; alcoholism; antithrombotic medications (warfarin, acetylsalicylic acid, cilostazol, clopidogrel, prasugrel, ticagrelor, ticlopidine, dabigatran etexilate, apixaban, edoxaban, rivaroxaban) tricyclic antidepressant; other AD use | NA |
| *SELF-CONTROLLED STUDIES* | | | |
| Rhode, 2022 [27] | Mean percent change | Individuals serve as their own controls, allowing adjustment for all time-stable confounders; findings externally compared to those from an age-sex matched reference population | NA |
| CASE-CONTROL STUDIES | | | |
| Noordam, 2016 [29] | Conditional logistic regression | Age; sex; BMI; number of prescribed oral GLDs | NA |
| *CROSS-SECTIONAL STUDIES* | | | |
| Higgins, 2007 [30] | Logistic regression | None for relevant analyses | Coronary artery disease, myocardial infarction, angioplasty, Coronary artery bypass graft |
| Yekta, 2015 [33] | Logistic regression | Age; sex; ethnicity; income; diabetes duration; HbA1c; SBP; TG levels; BMI; smoking | NA |
| Kammer, 2016 [31] | Linear regression | Age; sex; ethnicity; education; depression severity; BMI; smoking | NA |

Supplemental Table 4 **(b)** Additional information on adjustment for confounding in statistical models for the association between antidepressant prescribing and outcomes

| First author, year | Statistical methods | Confounders adjusted for in analyses | Details of composite outcomes (where relevant) |
| --- | --- | --- | --- |
| *COHORT STUDIES* | | |  |
| Spoelstra, 2004 [23] | Cox proportional hazards | Age at diabetes onset (first prescription of oral GLD); other drug prescriptions in previous 180 days^§^; calendar year | NA |
| Wu, 2016 [24] | Cox proportional hazards | Age at diabetes diagnosis; sex; calendar year of diabetes diagnosis; comorbid diagnoses^¶^; other drug prescriptions^#^; number of/adherence to prescribed oral GLDs; insulin prescription; examination for HbA1c/lipids; number of outpatient visits/hospitalisations | CV morbidity (hospitalisation for CHD and stroke; PVD with stent insertion; vascular shunt or bypass; vessel repair procedure), microvascular morbidity (retinopathy; blindness; end-stage renal disease with dialysis; vessel operations for haemodialysis; kidney transplantation; hospitalisation for diabetic foot infection; lower extremity amputations), all-cause mortality |
| *CASE-CONTROL STUDIES* | | |  |
| Lipscombe, 2009 [28] | Conditional logistic regression | Age; sex; neighbourhood income quintile; diabetes duration; CCI; comorbid dementia, schizophrenia, or other major psychoses; other drug prescriptions in previous 180 days**; health service use history | NA |
| *CROSS-SECTIONAL STUDIES* | | |  |
| Wake, 2016 [32] | Student’s t-test/chi-squared test | Exposed and non-exposed people, matched for: birth year; sex; diabetes type; diagnosis date; BMI; smoking | NA |

Supplementary Table 5 (a): Supplementary results of observational studies reporting on the association between antidepressant drug prescribing and outcomes in people with diabetes

| First author, year | Statistical methods | Potential confounding factors adjusted for | Results | | | | |
| --- | --- | --- | --- | --- | --- | --- | --- |
|  |  |  | Outcome | Reference | Antidepressant prescription | Effect estimate (CI)^†^ | Direction of association^‡^ |
| Rubin, 2010, 2013 [22] | Generalised estimating equations | Age; sex; ethnicity; education; outcome of interest in prior year; history of CVD; duration of diabetes; AD prescription (prior year); year of follow-up | Sub-optimal glycaemic control (HbA1c > 7%/53 mmol/mol) or insulin prescription | No AD | AD in prior year | *Active arm:* OR 1.25 (1.08-1.46)* | ↑ |
|  |  |  | SBP ≥ 130 mmHg or anti-hypertensive prescription | No AD | AD in prior year | *Control arm:* OR 1.09 (0.87-1.36) | ↔ |
|  |  |  |  |  |  | *Active arm:* OR 1.18 (0.94-1.48) | ↔ |
|  |  |  | DBP ≥ 80 mmHg or anti-hypertensive prescription | No AD | AD in prior year | *Control arm:* OR 1.06 (0.85-1.33) | ↔ |
|  |  |  |  |  |  | *Active arm:* OR 1.39 (1.11-1.74)* | ↑ |
|  |  |  | LDL ≥ 2.6 mmol/l or lipid-lowering prescription | No AD | AD in prior year | *Control arm:* OR 1.24 (0.96-1.60) | ↔ |
|  |  |  |  |  |  | *Active arm:* OR 1.15 (0.90-1.46) | ↔ |
|  |  |  | HDL ≤ 1.0 mmol/l  or lipid-lowering prescription | No AD | AD in prior year | *Control arm:* OR 1.24 (1.03-1.50)* | ↑ |
|  |  |  |  |  |  | *Active arm:* OR 1.33 (1.11-1.58)* | ↑ |
|  |  |  | TC ≥ 5.2 mmol/l or lipid-lowering prescription | No AD | AD in prior year | *Control arm:* OR 1.29 (1.05-1.57)* | ↑ |
|  |  |  |  |  |  | *Active arm:* OR 1.21 (1.00-1.48) | ↔ |
|  |  |  | TG ≥ 1.7 mmol/l or lipid-lowering prescription | No AD | AD in prior year | *Control arm:* OR 1.23 (0.99-1.52) | ↔ |
|  |  |  |  |  |  | *Active arm:* OR 1.75 (1.43-2.14)* | ↑ |
| Würtz, 2016 [26] | Cox proportional hazards | Multivariable adjusted/propensity score matched: previous MI, atrial ﬁbrillation or ﬂutter; intermittent arterial claudication; dementia; CCI (excluding MI, PVD, and dementia); other drug prescriptions^§^ | Mortality (30 days following any stroke)^\|\|^ | No SSRI | Current SSRI (any) | *Propensity score matched: RR* 1.2 (1.0-1.4) | ↔ |
|  |  |  |  |  | New SSRI | *Propensity score matched: RR* 1.4 (1.0-1.9) | ↔ |
|  |  |  |  |  | Long-term SSRI | *Propensity score matched: RR* 1.3 (1.1-1.6)* | ↑ |
|  |  |  |  |  | Former SSRI | *Propensity score matched: RR* 1.2 (0.8-1.7) | ↔ |
|  |  |  | Mortality (30 days following ischaemic stroke) | No SSRI | Current SSRI | Rate ratio 1.3 (1.1-1.7)* | ↑ |
|  |  |  |  |  |  | *Propensity score matched: RR* 1.4 (1.0-1.8) | ↔ |
|  |  |  |  |  |  | *Men:* RR 1.6 (1.2-2.2)* | ↑ |
|  |  |  |  |  |  | *Women:* RR1.2 (0.9-1.6) | ↔ |
|  |  |  |  |  |  | *Age < 60:* RR 1.8 (0.7-4.5) | ↔ |
|  |  |  |  |  |  | *Age 60 to 69:* RR 1.5 (0.9-2.8) | ↔ |
|  |  |  |  |  |  | *Age 70 to 79:* RR 1.6 (1.1-2.4)* | ↑ |
|  |  |  |  |  |  | *Age ≥ 80:* RR 1.2 (0.9-1.6) | ↔ |
|  |  |  | Mortality (30 days following intracerebral haemorrhage) | No SSRI | Current SSRI | RR 1.2 (0.9-1.5) | ↔ |
|  |  |  |  |  |  | *Propensity score matched:* RR 1.0 (0.7-1.3) | ↔ |
|  |  |  |  |  |  | *Men:* RR 1.1 (0.7 to 1.5) | ↔ |
|  |  |  |  |  |  | *Women:* RR 1.3 (0.9-1.9) | ↔ |
|  |  |  |  |  |  | *Age < 60:* RR 0.7 (0.2-2.3) | ↔ |
|  |  |  |  |  |  | *Age 60 to 69:* RR 1.3 (0.7-2.5) | ↔ |
|  |  |  |  |  |  | *Age 70 to 79:* RR 1.3 (0.8-2.2) | ↔ |
|  |  |  |  |  |  | *Age ≥ 80:* RR 1.4 (0.9- 2.0) | ↔ |
|  |  |  | Mortality (30 days following subarachnoid haemorrhage)^\|\|^ | No SSRI | Current SSRI | RR 0.8 (0.4-1.7) | ↔ |
|  |  |  |  |  |  | *Propensity score matched:* RR 0.9 (0.4-2.0) | ↔ |
|  |  |  | Mortality (30 days following unspecified stroke)^\|\|^ | No SSRI | Current SSRI | RR 1.2 (1.0-1.4) | ↔ |
|  |  |  |  |  |  | *Propensity score matched:* RR 1.2 (1.0-1.6) | ↔ |
| Hazuda, 2019^#^ [19] | Cox proportional hazards | Age; ethnicity; history of CVD; HbA1c; BMI; waist circumference; insulin prescription; hypercholesterolaemia; hypertension; smoking; estimated exercise stress level; HRT prescription (women only); clinic attended; assigned clinical trial intervention | CV mortality, non-fatal MI, non-fatal stroke | No AD | Baseline AD | *Men:* HR 0.72 (0.50-1.05) | ↔ |
|  |  |  |  |  |  | *Women:* HR 0.86 (0.61-1.21) | ↔ |
|  |  |  | All-cause mortality, non-fatal MI, non-fatal stroke, angina | No AD | Baseline AD | *Men:* HR 1.03 (0.80-1.34) | ↔ |
|  |  |  |  |  |  | *Women:* HR 0.72 (0.56-0.94)* | ↓ |
|  |  |  | CV mortality, non-fatal MI, non-fatal stroke, angina, CHF, PVD, CABG, carotid endarterectomy | No AD | Baseline AD | *Men:* HR 1.07 (0.85-1.36) | ↔ |
|  |  |  |  |  |  | *Women:* HR 0.76 (0.59-0.96)* | ↓ |
| Rohde, 2021 [21] | Cox proportional hazards | Age; sex; marital status; HbA1c (baseline); LDL levels (baseline); obesity; kidney functioning; CCI (excluding diabetes); diabetes complications; alcohol-related disorders; smoking-associated disorders; other drug prescriptions^¶^ | Optimal glycaemic control (HbA1c < 7%/53 mmol/mol) | No AD | Current SSRI | OR 0.94 (0.87-1.02) | ↔ |
|  |  |  |  |  | Current SNRI | OR 1.30 (1.10-1.52)* | ↓ (of sub-optimal glycaemic control) |
|  |  |  |  |  | Current TCA | OR 0.99 (0.85-1.15) | ↔ |
|  |  |  |  |  | Current other AD | OR 0.98 (0.86-1.12) | ↔ |
|  |  |  |  |  | Recent AD^#^ | OR 1.02 (0.96-1.08) | ↔ |
|  |  |  |  |  | Persistent AD^#^ | OR 1.00 (0.95-1.06) | ↔ |
|  |  |  | LDL < 2.6 mmol/l | No AD | Current AD | OR 1.08 (1.03-1.14)* | ↓ (of sub-optimal LDL) |
|  |  |  |  |  | Current SSRI | OR 1.04 (0.98-1.12) | ↔ |
|  |  |  |  |  | Current SNRI | OR 1.26 (1.12-1.42)* | ↓ (of sub-optimal LDL) |
|  |  |  |  |  | Current TCA | OR 1.22 (1.08-1.38)* | ↓ (of sub-optimal LDL) |
|  |  |  |  |  | Other AD | OR 0.99 (0.89-1.11) | ↔ |
|  |  |  |  |  | Former AD | OR 1.06 (1.01-1.11)* | ↓ (of sub-optimal LDL) |
|  |  |  |  |  | Recent AD^#^ | OR 1.05 (1.00-1.10) | ↔ |
|  |  |  |  |  | Persistent AD^#^ | OR 1.06 (1.01-1.11)* | ↓ (of sub-optimal LDL) |
|  |  |  | GLD prescription (including insulin) | No AD | Current AD (sub-cohort)** | OR 1.34 (1.29-1.39)* | ↑ |
|  |  |  |  |  | Current SSRI | OR 1.28 (1.22-1.34)* | ↑ |
|  |  |  |  |  | Current SNRI | OR 1.76 (1.63-1.89)* | ↑ |
|  |  |  |  |  | Current TCA | OR 1.34 (1.23-1.46)* | ↑ |
|  |  |  |  |  | Current other AD | OR 1.45 (1.35-1.56)* | ↑ |
|  |  |  |  |  | Recent AD^#^ | OR 1.37 (1.32-1.41)* | ↑ |
|  |  |  |  |  | Persistent AD^#^ | OR 1.38 (1.33-1.42)* | ↑ |
| Chen, 2021 [17] | Cox proportional hazards | Age; sex; income; urbanization; hypertension; hypercholesterolaemia; CAD; CKD; heart failure; peptic ulcer; aspirin; plavix | First MI | No AD | TCA (>180 days)  SSRI (>180 days)  SNRI (>180 days)  AD cDDD:  28-180  >180 | HR 0.74 (0.69-0.79)*  HR 0.66 (0.60- 0.74)*  HR 0.67 (0.51- 0.88)*  HR 0.77 (0.73-0.81)  HR 0.56 (0.52-0.60) | ↓  ↓  ↓  ↓  ↓ |
| Wu, 2021 [25] | Cox proportional hazards | Only crude incidence rates reported (adjusted models only reported for categories of use versus “poor use”) | Macrovascular complications  Microvascular complication  All-cause mortality | No AD | SSRIs  SNRIs  TCAs  SSRIs  SNRIs  TCAs  SSRIs  SNRIs  TCAs | 69.5 vs 65.6 per 1000 person-years  75.4 vs 65.6 per 1000 person-years  98.8 vs 65.6 per 1000 person-years  41.7 vs 40.9 per 1000 person-years  51.8 vs 40.9 per 1000 person-years  48.4 versus 40.9 per 1000 person-years  18.3 vs 17.3 per 1000 person-years  19.5 vs 17.3 per 1000 person-years  26.9 vs 17.3 per 1000 person-years | ↑^††^  ↑^††^  ↑^††^  ↑^††^  ↑^††^  ↑^††^  ↑^††^  ↑^††^  ↑^††^ |
| Chen, 2022 [18] | Cox proportional hazards | Age; sex; economic level; urbanization level; CCI score; hypertension; acute MI, hyperlipidemia; atrial fibrillation; COPD; depression; bipolar disorder; schizophrenia; alcoholism; antithrombotic medications (warfarin, acetylsalicylic acid, cilostazol, clopidogrel, prasugrel, ticagrelor, ticlopidine, dabigatran etexilate, apixaban, edoxaban, rivaroxaban) tricyclic antidepressant; other AD use | PVD | No SSRI | SSRI average dose effects:  cDDD: 1-83  cDDD: ≥84 | HR 1.17 (0.74-1.83)  HR 1.04 (0.50-2.15) | ↔  ↔ |

*Denotes significant P < 0.05

^†^Results given for fully multivariable adjusted or matched models unless otherwise stated

^‡^No association refers to no statistically significant association at P < 0.05

^§^Including: angiotensin system acting agents; beta blockers, calcium channel blockers; statins; aspirin; non-aspirin non-steroidal anti-inflammatory drugs; non-aspirin platelet inhibitors; vitamin K antagonists; corticosteroids for systemic use; APs

^||^Results not given for age or sex stratified analyses

^¶^Including: angiotensin system acting agents; beta blockers; calcium channel blockers; diuretics; corticosteroids for systemic use; antithrombotic agents; analgesics; inhalants

^#^Recent AD prescription defined as prescription 101 to 365 days prior to the diagnosis of T2DM, persistent AD prescription defined as ≥ 2 prescriptions in the 2 years prior to the diagnosis of T2DM including a prescription in the year prior to the diagnosis of T2DM

**Restricted to people diagnosed with T2DM after 2007 to account for differences in treatment guidelines

^††^Crude incidence rates reported – unadjusted for age and sex

Abbreviations: AD: antidepressant; AP: antipsychotic; BMI: body mass index; CABG: coronary artery bypass graft; CCI: Charlson Comorbidity Index; CHF; congestive heart failure; CI: confidence interval; CV: cardiovascular; CVD: cardiovascular disease; DBP: diastolic blood pressure; GLD: glucose lowering drug; HbA1c: glycated haemoglobin; HDL: high-density lipoprotein; HR: hazard ratio; HRT: hormone replacement therapy; LDL: low-density lipoprotein; MI: myocardial infarction; OR: odds ratio; PVD: peripheral vascular disease; RR; rate ratio; SBP: systolic blood pressure; SNRI: serotonin–norepinephrine reuptake inhibitor; SSRI: selective serotonin reuptake inhibitor; T2DM: type 2 diabetes mellitus; TC: total cholesterol; TCA: tricyclic antidepressant; TG: triglyceride

### Supplemental Table 5 (b): Supplementary results of observational studies reporting on the association between antipsychotic drug prescribing and outcomes in people with diabetes

| First author, year | Statistical methods | Potential confounding factors adjusted for | Results | | | | |
| --- | --- | --- | --- | --- | --- | --- | --- |
|  |  |  | Outcome | Reference | Antipsychotic prescription | Effect estimate (CI)^†^ | Direction of association^‡^ |
| Wu, 2016 [24] | Cox proportional hazards^§^ | Multivariable adjusted/propensity score matched: age at diabetes diagnosis; sex; calendar year of diabetes diagnosis; comorbid diagnoses^\|\|^; other drug prescriptions^¶^; number of/adherence to prescribed GLDs; insulin prescription; examination for HbA1c/lipids; number of outpatient visits/hospitalisations | All complications | No AP | Irregular AP | HR 0.90 (0.78 to 1.03) | ↔ |
|  |  |  |  |  | Regular AP (stratified by metabolic risk)^#^ | *Overall:* HR 0.81 (0.69 to 0.95)* | ↓ |
|  |  |  |  |  |  | *Overall, propensity score matched:* HR 0.75 (0.67 to 0.84)* | ↓ |
|  |  |  |  |  |  | *Low:* HR 0.85 (0.70 to 1.02) | ↔ |
|  |  |  |  |  |  | *Intermediate:* HR 0.82 (0.68 to 0.99)* | ↓ |
|  |  |  |  |  |  | *High:* HR 0.69 (0.53 to 0.91)* | ↓ |
|  |  |  |  |  |  | *Combination:* HR 0.84 (0.66 to 1.08) | ↔ |
|  |  |  | CV morbidity | No AP | Regular AP (stratified by metabolic risk)^#^ | *Overall, propensity score matched:* HR 0.71 (0.62 to 0.80)* | ↓ |
|  |  |  |  |  |  | *Low:* HR 0.84 (0.67 to 1.06) | ↔ |
|  |  |  |  |  |  | *Intermediate:* HR 0.81 (0.64 to 1.02) | ↔ |
|  |  |  |  |  |  | *High:* 0.74 (0.53 to 1.02) | ↔ |
|  |  |  |  |  |  | *Combination:* HR 0.75 (0.55 to 1.04) | ↔ |
|  |  |  | Microvascular morbidity | No AP | Regular AP (stratified by metabolic risk)^#^ | *Overall, propensity score matched:* HR 0.87 (0.73 to 1.03) | ↔ |
|  |  |  |  |  |  | *Low:* HR 0.83 (0.62 to 1.11) | ↔ |
|  |  |  |  |  |  | *Intermediate:* HR 0.83 (0.62 to 1.10) | ↔ |
|  |  |  |  |  |  | *High:* HR 0.61 (0.40 to 0.93)* | ↓ |
|  |  |  |  |  |  | *Combination:* HR 1.03 (0.72 to 1.46) | ↔ |
|  |  |  | All-cause mortality | No AP | Regular AP (stratified by metabolic risk)^#^ | *Overall, propensity score matched*: HR 0.79 (0.71 to 0.89)* | ↓ |
|  |  |  |  |  |  | *Low:* HR 0.66 (0.54 to 0.81)* | ↓ |
|  |  |  |  |  |  | *Intermediate:* HR 0.78 (0.65 to 0.94)* | ↓ |
|  |  |  |  |  |  | *High:* HR 0.62 (0.47 to 0.82)* | ↓ |
|  |  |  |  |  |  | *Combination:* HR 0.82 (0.65 to 1.05) | ↔ |
| Lipscombe 2009 [28] | Conditional logistic regression | Age; sex; neighbourhood income quintile; diabetes duration; CCI; comorbid dementia, schizophrenia, or other major psychoses; other drug prescriptions in previous 180 days**; health service use history | Hospitalisation for hyperglycaemia | Remote AP | Current FGA (any) | *Insulin:* rate ratio 1.27 (0.75 to 2.12) | ↔ |
|  |  |  |  |  |  | *Oral GLD:* rate ratio 1.31 (0.90 to 1.90) | ↔ |
|  |  |  |  |  |  | *None:* rate ratio 3.43 (1.59 to 7.38)* | ↑ |
|  |  |  |  |  | Current SGA (any) | *Insulin:* rate ratio 1.40 (1.06 to 1.85)* | ↑ |
|  |  |  |  |  |  | *Oral GLD:* rate ratio 1.37 (1.12 to 1.67)* | ↑ |
|  |  |  |  |  |  | *None:* rate ratio 2.37 (1.57 to 3.58)* | ↑ |
| Wake, 2016 [32] | Student’s t-test/chi-squared test | Exposed and non-exposed people, matched for: birth year; sex; diabetes type; diagnosis date; BMI; smoking | Mean SBP (mmHg) ± SD | AP (≥ 12 months in total) | | 130.7 ± 2.0* | ↓ |
|  |  |  |  | No AP | | 134.5 ± 1.1 |  |
|  |  |  | Mean DBP (mmHg) ± SD | AP (≥ 12 months in total) | | 74.3 ± 1.2* | ↓ |
|  |  |  |  | No AP | | 75.1 ± 0.7 |  |
|  |  |  | Mean TC (mmol/l) ± SD | AP (≥ 12 months in total) | | 4.2 ± 0.2* | ↓ |
|  |  |  |  | No AP | | 4.3 ± 0.1 |  |

*Denotes significant P < 0.05

^†^Results given for fully multivariable adjusted or matched models unless otherwise stated

^‡^No association refers to no statistically significant association at P < 0.05

^§^Time-dependent with AP prescribing measured in six month intervals

^||^Including: hypertension; dyslipidaemia; chronic pulmonary disease; chronic liver disease; malignancy; depression; dementia; anxiety disorders; alcohol-related disorders; substance use disorders

^¶^Including: angiotensin system acting agents; beta blockers; calcium channel blockers; diuretics; lipid-lowering agents; antithrombotic agents; non-steroidal anti-inflammatory drugs; anticonvulsants; lithium; ADs; benzodiazepines

^#^Drugs considered to have high metabolic risk included: clozapine; olanzapine, drugs considered to have intermediate metabolic risk included: paliperidone; quetiapine; risperidone; zotepine; chlorpromazine; chlorprothixene; clopentixol; clothiapine; loxapine; methotrimeprazine; perphenazine; pipotiazine; prochlorperazine; thioridazine; zuclopentixol, drugs considered to have low metabolic risk included: amisulpride; aripiprazole; sulpiride; ziprasidone; flupentixol; fluphenazine; haloperidol; pimozide; thiothixene; trifluoperazine

**Including: cytochrome P-450 2C9 inducers; cytochrome P-450 2C9 inhibitors; thiazide diuretics; corticosteroids for systemic use

Abbreviations: AD: antidepressant; AP: antipsychotic; BMI: body mass index; CCI: Charlson Comorbidity Index; CI: confidence interval; CV: cardiovascular; DBP: diastolic blood pressure; FGA: first-generation antipsychotic; GLD: glucose lowering drug; HbA1c: glycated haemoglobin; HR: hazard ratio; SBP: systolic blood pressure; SGA: second-generation antipsychotic; TC: total cholesterol
